# Archipelago method for variant set association test statistics

**DOI:** 10.1101/2025.03.17.25324111

**Authors:** Dylan Lawless, Ali Saadat, Mariam Ait Oumelloul, Luregn J. Schlapbach, Jacques Fellay

## Abstract

Variant set association tests (VSAT), especially those incorporating rare variants via variant collapse, are invaluable in genetic studies. However, unlike Manhattan plots for single-variant tests, VSAT statistics lack intrinsic genomic coordinates, hindering visual interpretation. To overcome this, we developed the Archipelago method, which assigns a meaningful genomic coordinate to VSAT P values so that both set-level and individual variant associations can be visualised together. This results in an intuitive and information rich illustration akin to an Archipelago of clustered islands, enhancing the understanding of both collective and individual impacts of variants. We conducted three validation studies spanning simulated and real datasets across small and biobank-scale cohorts, from 504 individuals up to 490,640 UK Biobank participants. We integrated single-variant genome-wide association studies (GWAS) with gene-and protein pathway-level rare-variant collapse. These studies included the 1KG GWAS cohort, the Pan-UK Biobank GWAS with DeepRVAT WES gene-level study, and the UKBB WGS gene-level UTR collapsing PheWAS. The Archipelago plot is applicable in any genetic association study that uses variant collapse to evaluate both individual variants and variant sets, and its customisability facilitates clear communication of complex genetic data. By integrating at least two dimensions of genetic data into a single visualisation, VSAT results can be easily read and aid in identification of potential causal variants in variant sets such as protein pathways.

**GitHub repository:** https://github.com/DylanLawless/archipelago.

**CRAN submission:** Archipelago Version 0.0.1.9000.

**Zenodo:** https://doi.org/10.5281/zenodo.16880622.

## 1 Introduction

Variant set association tests (VSATs) are methods in which groups of variants are collapsed and analysed jointly to enhance an association signal. VSAT, particularly those incorporating rare variants (e.g. minor allele frequencies below 1%), have become indispensable in genetic association studies (1–6). Next-generation sequencing has enabled the comprehensive detection of rare variants, thereby complementing traditional single-variant tests (7; 8). While Manhattan plots effectively visualise single-variant associations (9), a comparable graphical representation for VSAT results has been lacking.

To name a small few, VSAT methods include burden tests (e.g. CAST (10), CMC (11), weighted-sum (12)), adaptive burden tests (e.g. alpha-sum (13), VT (14), KBAC (15)) and variance component tests. Variance component approaches, such as the C(*α*) test (16; 17) and kernel machine regression methods (3), offer flexible strategies for assessing the combined effect of rare variants. In particular, the sequence kernel association test (SKAT) (18) and its extensions including SKAT-O (19), MSKAT (20), famSKAT, and RC-SKAT (21–24) have become widely adopted.

More recently, the variant-set test for association using annotation information (STAAR) framework (5; 25) has been introduced, utilising the aggregated Cauchy association test (ACAT) (26) to combine multiple annotation-based P values for each variant. Despite these methodological advances, the lack of a natural genomic coordinate for VSAT P values hinders their visual interpretation.

To address this gap, we introduce the Archipelago plot. By assigning a genomic coordinate to the VSAT P value based on the average of its constituent variants, this method facilitates the simultaneous visualisation of aggregated and individual variant effects. Notably, multiple types of genome-wide association study (GWAS), VSAT, and rare variant association test (RVAT), including SKAT-O and ACAT, have been highly successful and scalable to national biobank cohorts where this method can be applied. Examples include a Pan-UK Biobank (UKBB) GWAS (27) of 7,266 phenotypes (~441k individuals), DeepRVAT gene-level rare variant WES analyses (~470k individuals) (28), the UKBB WGS UTR collapsing PheWAS (~490k individuals) (8). Meta-analysis of TOPMed whole-genome sequencing data and UKBB whole-exome sequencing data, encompassing ~200k individuals, has demonstrated the utility of these methods (29). Similarly, rare non-coding variation in complex human phenotypes has been investigated on ~333k individuals from three cohorts: UK Biobank (N = ~200k), TOPMed (N = ~88k) and All of Us (N = ~45k) (30). Given their proven success, VSAT methods are likely to remain essential in future studies, and our Archipelago plot is designed to be applicable to such investigations.

## 2 Implementation

The Archipelago plot is a novel visualisation technique developed to facilitate the interpretation of both (1) variant-set association testing (VSAT) P values (2) single variant P values in the context of the variant set. Its design is based on the commonly used Manhattan plot but accounts for the unique properties of VSAT statistics. **Figures 1** uses synthetic data to illustrate output that might be expected in RVAT for case-control analysis. Briefly, 5,000 independent variants were simulated for GWAS results and 250 VSAT tests (20 variants per set). VSAT P values were drawn from a log-normal distribution using a minimal threshold (0.05/10), while GWAS P values were uniformly sampled between 0.05/100 and 1, with base pair positions and chromosome numbers assigned at random. This represented 250 VSAT P values and 5000 independent variants. As shown in **figure 1**, blue points indicate the joint VSAT P value as produced by SKAT-O or other statistical methods. Alternating yellow and orange points indicate the individual P values from each variant as produced by a regression or some other statistical method (e.g. R package SKAT: `$param$p.val.each`, P value for each single variant in a set-based test, or as calculated in single variant analysis). Alternating colours clarify the chromosome position. Lines connect the VSAT P value (blue) to its constituent individual variants to indicate variant set grouping. These edges are only shown for variants which are also in the significantly enriched VSAT P value group.

**Figure 1:**
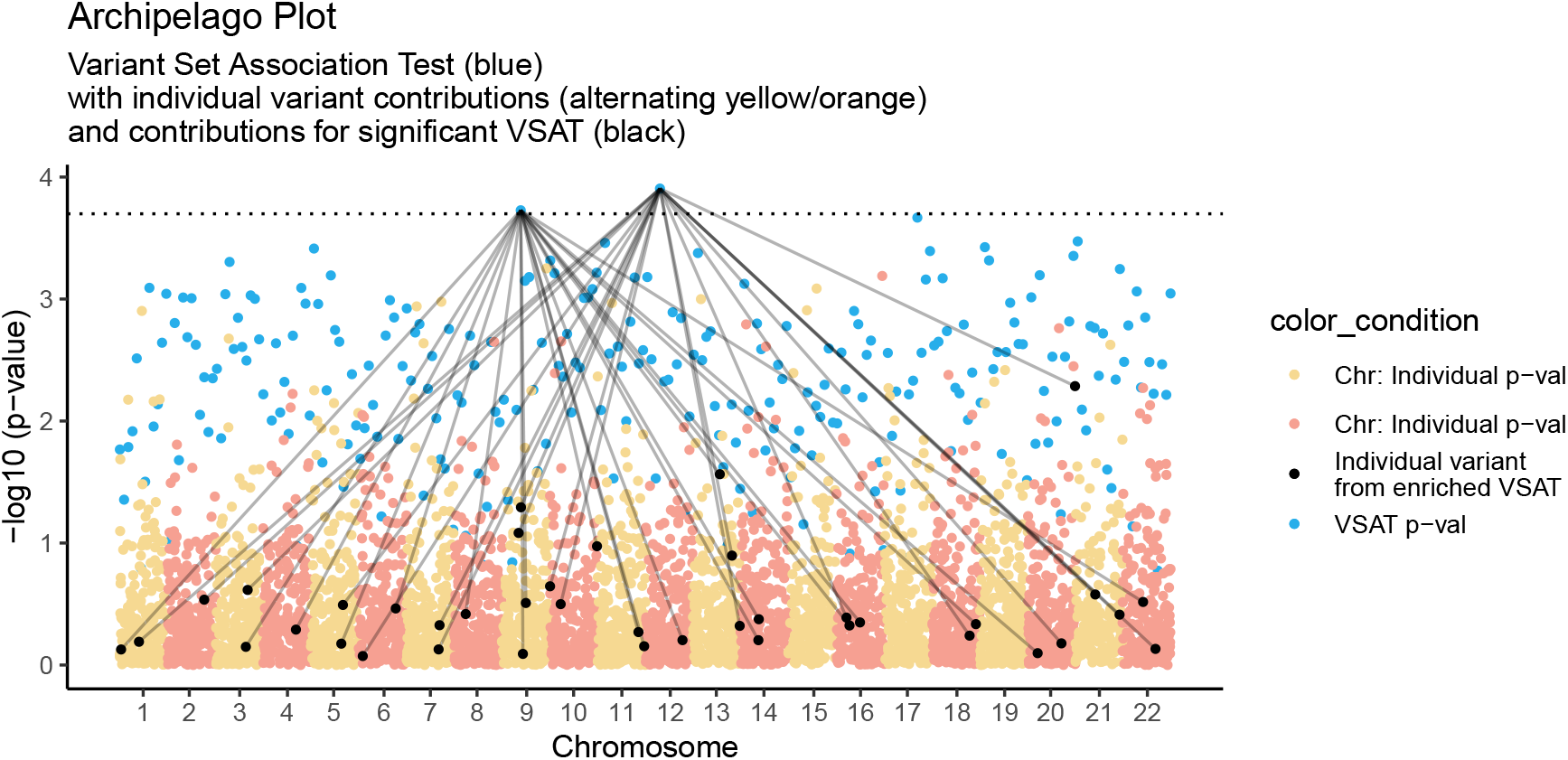
Synthetic data for human autosomal chromosomes 1-22. Contains 250 variant sets with 5000 individual variants. Each significantly enriched VSAT P value (blue) is mapped to its constituent individual P values (yellow and orange points). Significance threshold based on the number of VSATs.

The precise protocol summary, definition, and algorithm can be found in **sections 13.7–13.9**. User settings of the R package can be read in **section 13.6**. The R code used in this manuscript can be found in the GitHub package repository. The main steps of the process are:

1. Within each variant set, get the average genome-wide genomic coordinate and assign it to the VSAT group P value.
2. Optionally, normalise the VSAT x-axis distribution for dispersion to prevent centre clustering in dense datasets. Ranking of VSAT P values also provides priority in the event of overlaps.
3. Optionally, map the VSAT position to each individual variant from the set.

Since variant sets (such as protein pathways) have no specific single genomic coordinate, it is otherwise difficult to assign a logical x-axis position. Figure 2 illustrates the default format of raw VSAT P values, which will either be ranked by their P value strength or some other rank based on the variant set construction which results in an arbitrary x-axis distribution. The Archipelago plot assigns these same VSAT results into their x-axis location as the ranked average of the individual variant set genomic coordinate, thus representing an information-rich illustration. For example, a variant set that is made from mostly chromosome 1 variants, will appear on the left, near its constituent variant P value points from chromosome 1. Genome-wide variant sets will appear near the centre, however to prevent strong clustering in very dense datasets we also provide the method for ranked dispersion based on genomic coordinates for clearer illustration of the VSAT P value.

**Figure 2:**
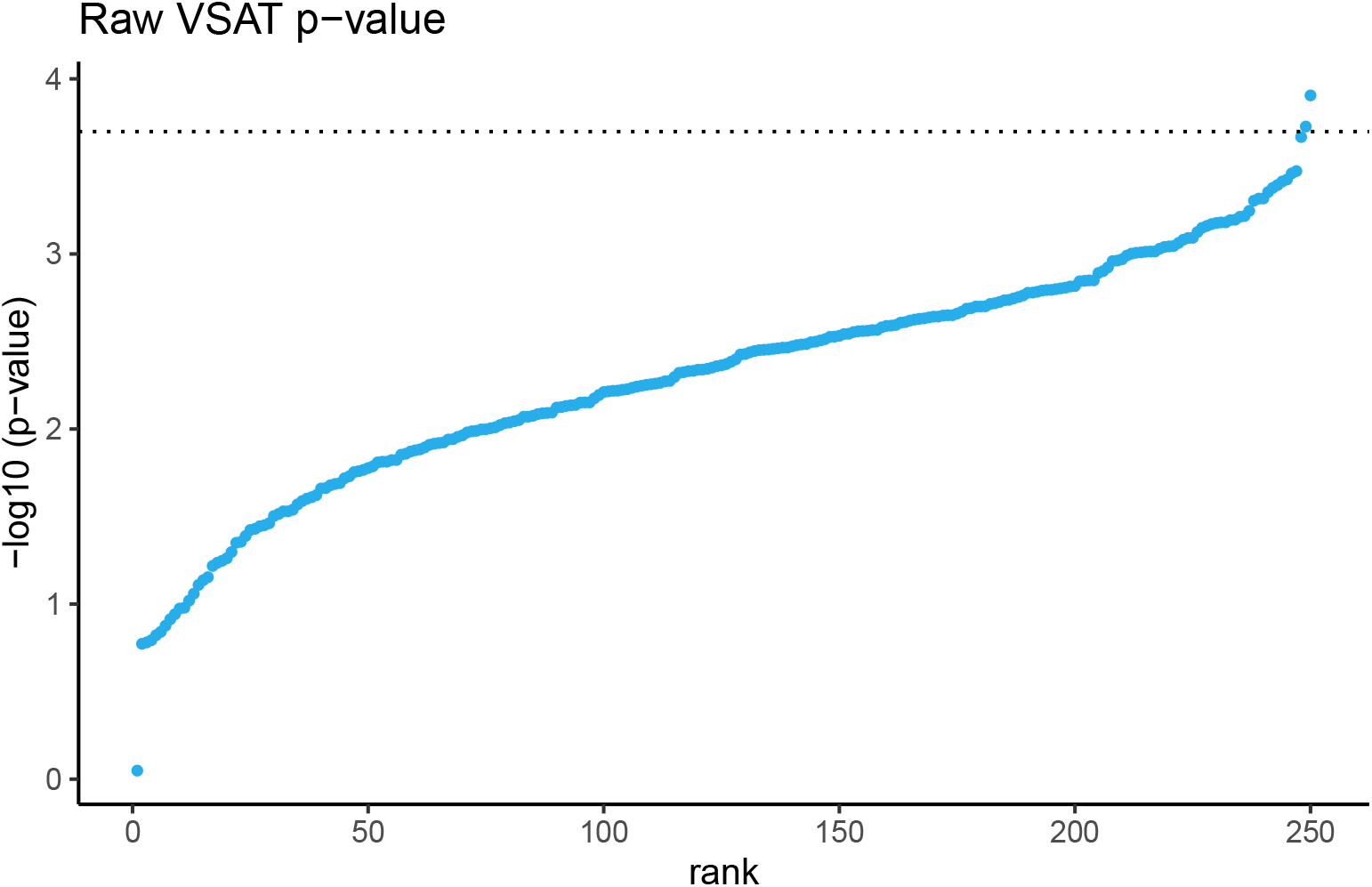
Raw VSAT P values have no natural x-axis position and must be ranked on association strength or some arbitrary ranking. The same VSAT result shown in **figure 1** (blue) is shown here without the use of the Archipelago method.

To ensure that VSAT points do not cluster and heavily overlap, ranking is applied before finding the variant set average based on the VSAT P value. That is, the x-axis position of the strongest association will have priority. Although overlap is unlikely in genome-wide testing, it may occur where variant sets are constructed from a narrow set of genomic locations. In all figures a significant P value threshold was used which would typically be derived based on the number of independent tests, such as the number of variant sets. In these examples we used the arbitrarily chosen 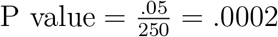 indicating 250 variant set tests.

We include a set of supplemental **figures S2a, S2b**, and **S2c** which contain the same dataset as **figure 1**. However, each version illustrates the decreasing levels of clarification information to demonstrate the layers of annotation. **Figure S2a** shows the original plot without the figure legend. **Figure S2b** drops the edge highlights for the significantly enriched VSAT to reveal all connections, which can be difficult to read in high density plots. **Figure S2c** next drops the individual variant P value highlighting. Conversely, **figure S3** adds an additional layer of information by adding two colours for the significantly enriched variant sets, which is useful when there are multiple enriched variant sets but otherwise may be distracting. We demonstrate a set of sparse plots in **figures S4a, S4b**, and **S4c**. These smaller dataset examples use synthetic data to represent 500 qualifying variants, and VSAT of 20 genes/variants per variant set (25 VSAT P values).

Customisation settings are described in section 13.10. **Figure S5** shows an example of customised colours. **Figure S6** shows all customisable elements (title, subtitle, colours, colour labels, critical threshold line, genomic coordinate, show title and subtitle, and show legend). **Figures S7 - S8** shows all 16 colour themes.

## 3 Expansion with additional information layers

The variant-set test for association using annotation information (STAAR) method has special features since each individual variant P value (pSTAAR) is itself made from an aggregated P value derived from the set of annotation weighted P value using the ACAT method (5; 25; 26). Therefore, a third level of information hierarchy is present. In the basic Archipelago plot we have two layers: (1) VSAT P value and (2) individual P value. The third pSTAAR level might be illustrated in a sub-plot which focuses on the significantly enriched VSAT group. This sub-plot is then free to show x-axis genomic coordinates, and y-axis P value which is used for the Cauchy distribution aggregation of *f* (*p*_*n*_).

To demonstrate the source of annotation based ACAT P values, **Figure 3** panel [1] shows the Archipelago plot with two significantly enriched variant sets (blue), 40 contributing individual variants (black). Panel [2] then shows the ACAT P values for 4 annotation layers for each one of the individual (black) variants. To clarify that each single variant position will have four annotation P values, panel [3] overlays line/group box to show the spread of ACAT P values for each single variant. We note that highly correlated annotation layers are likely since many prediction tools rely on population genetics frequency as an indicator of variant effect quantification. Therefore, it would be reasonable to also show a further plot of the correlation between the annotation layers. While this is a simplified and specific example, we imagine that other similar applications will exist for VSAT with third layers of information that consist of x-axis genomic coordinate and y-axis test statistic.

**Figure 3:**
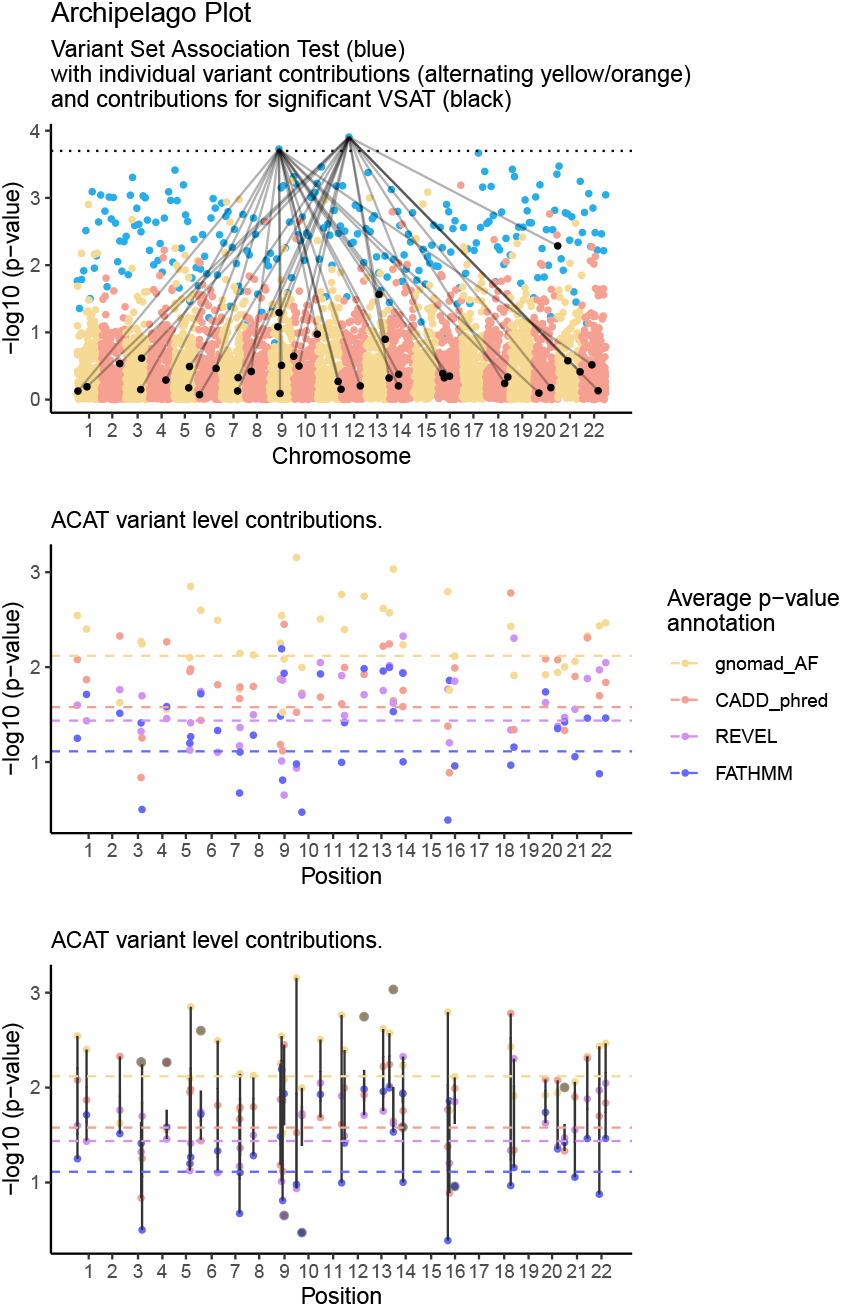
Panel [1] Archipelago plot as per Figure 1 with two significantly enriched variant sets (blue), 40 contributing individual variants (black). Panel [2] ACAT P values based on 4 annotation layers for each one of the individual (black) variants. Horizontal lines show the average annotation P value as an estimate of the hierarchy of contribution. Panel [3] overlays line/group box to show the spread of ACAT P values for each single variant.

## 4 Validation

We validated the Archipelago method across three distinct scenarios, spanning both simulated and real datasets of single-variant GWAS, and gene- or pathway-level collapse VSAT/RVAT:

1. **1000 Genomes (1KG)** - 504 East Asian samples with a simulated binary trait GWAS and a pathway-level VSAT (31).
2. **Pan-UK Biobank (UKBB)** - 469,382 UKBB samples with a quantitative trait GWAS and DeepRVAT gene-level WES RVAT (27; 28).
3. **UKBB WGS UTR PheWAS** - 490,640 UKBB samples with GWAS and untranslated region (UTR) collapsing gene-level WGS RVAT (8).

These settings demonstrate Archipelago’s versatility in unifying association signals across variant resolution and trait architecture.

### 4.1 Validation in 1KG using GWAS and simulated VSAT

We validated the Archipelago method by first reproducing a GWAS study (**supplemental, section 13**) using the public dataset of 504 East Asian individuals from the 1000 Genomes Project phase 3 (version 5, hg19) (31). Since 1KG data lack a common disease phenotype, we simulated a binary trait (250 cases, 254 controls) under a heritability of 0.8, then applied standard QC steps (e.g. splitting multi-allelic variants, normalising genotypes, LD pruning, KING-based relatedness checks) to retain 500 samples and 1,224,104 single nucleotide polymorphisms (SNPs). A single-variant GWAS was run via logistic regression with Firth correction, and a VSAT was conducted using protein pathways from ProteoMCLustR (941 pathways) and SKAT-O. **Tables S1–S2** illustrate the resulting style of pathway- and SNP-level outputs required; the Archipelago plot (**Figure 4 A**) integrates both in a single genomic view.

**Figure 4:**
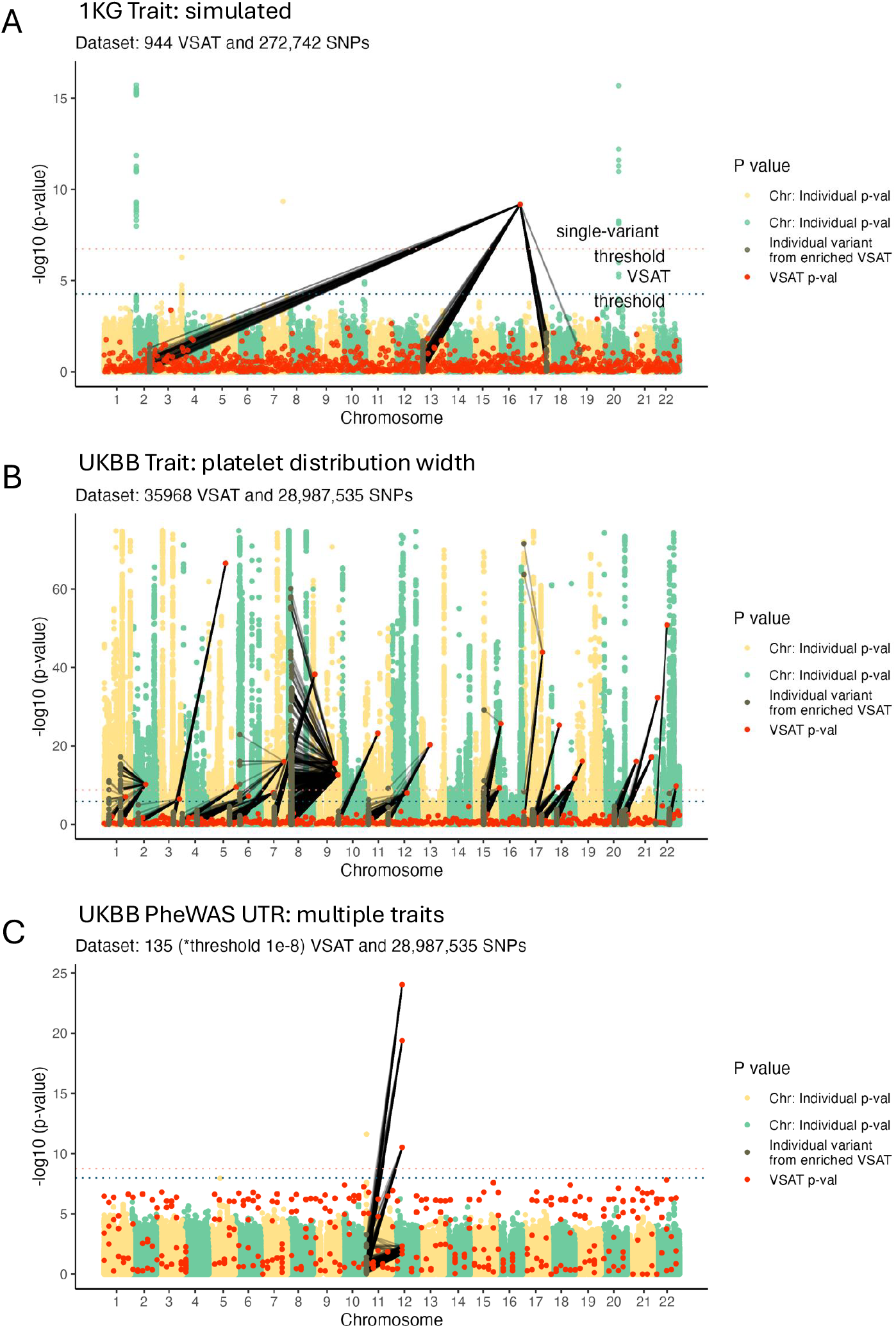
Archipelago plots across three validation settings. **A**: 1000 Genomes East Asian cohort (n=504) with simulated pathway-level binary trait (31). **B**: Pan-UK Biobank (n=469,382) platelet distribution width trait using WES GWAS and DeepRVAT gene-level VSAT (data trimmed at 1e-75 for distracting outliers) (27; 28). **C**: UKBB WGS UTR collapsing PheWAS (n=490,640) using WGS GWAS and UTR gene-level VSAT (*The UK Biobank Whole-Genome Sequencing Consortium et al. (8) defined the significance threshold for their PheWAS dataset) (8). In all panels, Archipelago visualises variant-level and set-level associations across the genome, enabling joint interpretation of individual and collapsed variant signals.

The enriched variant signals displayed in **Figure 4 A** were assessed based on their allele frequency and displayed in **Figure S1**. In this dataset, top GWAS signals involved highly common variants (n = 67, mean allele frequency 0.333, min = 0.13, max = 0.49), whereas the top VSAT signal (`set_ID` 532) included 261 variants with a mean frequency of 0.151 (min = 0.01, max = 0.48), reflecting a different genetic architecture captured by pathway-based collapse.

### 4.2 Validation in Pan-UK Biobank using GWAS and gene-level DeepRVAT

We selected a notable quantitative trait “platelet distribution width” for validation based on the large UKBB rare variant WES study (minor allele frequency <0.1%) for DeepRVAT by Clarke et al. (28). We performed the analysis using the two complementary layers: a Pan-UK Biobank SNP-level GWAS (27) and gene-level rare variant association results from DeepRVAT on UK Biobank WES (469,382 participants) (28). Together these layers comprise 29 million SNP-level GWAS statistics and 35,968 gene-level RVAT tests. The joint analysis result of Archipelago is shown in **Figure 4 (B)**.

For this highly polygenic and heritable trait, the Pan-UKBB GWAS identified hundreds of significant loci, including multiple regions with *P <* 10^−50^, consistent with known biology of platelet regulation. DeepRVAT identified 26 gene-level associations, capturing additional rare variant signals not resolved at the common single-variant level. The integrated view enabled systematic comparison of RVAT and GWAS signals, capturing convergence and divergence between rare and common variant associations. Eight genes (*APOA5, MOB3C, DOCK5, LPL, GP1BA, PEAR1, MPIG6B, PLEKHO2*) showed significant association in both GWAS and RVAT layers, revealing a novel insight.

### 4.3 Validation in UKBB WGS using GWAS and UTR collapsing PheWAS

We used the flagship UK Biobank whole-genome sequencing study of 490,640 participants as a large-scale, well characterised validation resource (8). Within this study, we selected the UTR gene-collapsing PheWAS as a notable analysis for integrating rare non-coding burden with GWAS, given its cross-ancestry design, deep phenotyping, and improved coverage of UTR and structural variation relative to whole exome sequencing (WES). A total of 135 traits were tested (e.g. ICD-10 D56 thalassaemia and ICD-10 I50 heart failure). The study reported a single genome-wide significant binary association in the UTR analysis, thalassaemia (ICD-10 D56), localising to *HBB* under the 5’+3’ UTR model at *P* = 9.16 *×* 10^−25^ with an empirically validated threshold of *P* ≤ 1 *×* 10^−8^ (figure 4 of The UK Biobank Whole-Genome Sequencing Consortium et al. (8)).

Using Archipelago, in **Figure 4 (C)** we reproduced *HBB* as the top binary UTR association and showed that this rare UTR burden maps back to individual *HBB* SNPs in the Pan-UKBB GWAS (27). Separately, the GWAS layer also exhibited a significant peak (yellow chromosome 11 SNPs) across the beta globin cluster on chr11p15.5, where the lead signal overlaps nearby genes including *HBE1, HBG2* and *OR51B5*, consistent with local linkage disequilibrium, while the UTR collapsing result attributes the gene-level effect to *HBB*. This validation provides a simple clinically interpretable positive control. Archipelago can thus illustrate set-level rare UTR burden to constituent GWAS variants and provide a novel insight using both layers.

## 5 Discussion

We have introduced the Archipelago plot as a novel method for interpreting VSAT results alongside individual variant P values, as produced from SKAT-O or some other statistical method after variant collapse (1; 2; 4). In contrast to traditional Manhattan plots that are restricted to single-variant associations with clear genomic coordinates, the Archipelago plot assigns a representative genomic location to the VSAT P value based on the average of its constituent variants (9). This enables a direct comparison of the aggregated signal with individual variant effects within a familiar framework.

The absence of a natural genomic coordinate for VSAT P values has previously hindered the integrated visual interpretation of variant set and single-variant results. By linking the VSAT P value to the genomic positions of the individual variants, the Archipelago plot offers an intuitive depiction of the relationship between variant set effects and the underlying individual signals. This approach may assist in identifying potential causal variants or pathways and can be readily extended to accommodate additional layers of information, such as annotation-based P values derived using methods like ACAT. We have seen that this method could be applied to national biobank scale genomic studies with relative ease (29; 30).

While the method was developed with rare-variant testing in mind, its customisable design permits application to any genetic association study that utilises variant collapse. Overall, the Archipelago plot provides a modest yet useful tool for the clear communication of complex genetic data, contributing to improved interpretability in large-scale VSAT analyses.

## 6 Author contributions

Dylan Lawless designed and wrote the work and performed analysis. Ali Saadat and Mariam Ait Oumelloul wrote the work. Luregn J. Schlapbach and Jacques Fellay led project management and funding.

## 9 Acknowledgements

We would like to thank all the patients and families who have been providing advice on SwissPedHealth and its projects, as well as the clinical and research teams at the participating institutions. We thank the 1KG, DeepRVAT, UKBB, and Pan-UKBB studies for their work.

## 10 Data availability

- This study is reproducible using the provided code repository https://github.com/DylanLawless/archipelago.
- The data repository additionally provides the validation studies data (1.5G), which are also reproducibly self-contained (32) https://doi.org/10.5281/zenodo.16880622.
- Detailed data sources and automated download links are provided within each script of the online data repository for the 1KG, Pan-UKBB, and DeepRVAT studies. The original sources are:
- The 1000 Genomes Project phase 3 version 5 (1KG 3v5), genome build human_g1k_v37.fasta (hg19) (31) http://ftp.1000genomes.ebi.ac.uk/vol1/ftp/release/20130502/.
- Pan-UK Biobank GWAS summary statistics: the UKBB GWAS (continuous-30110-both_sexes-irnt.tsv.bgz) and phenotype manifest (27) https://pan.ukbb.broadinstitute.org.
- DeepRVAT gene-trait association testing results on the 470k UK Biobank WES dataset (UKBB_470k_deeprvat_results.csv) (6; 28) https://doi.org/10.5281/zenodo.12736824.
- UKBB WGS UTR collapsing PheWAS results in 490k: Supplementary Table 16 from (8) (Significant and suggestive gene-phenotype associations identified in the UTR PheWAS collapsing analysis across 5’ UTR, 3’ UTR, 5’ + 3’ UTR and coding sequence + 5’ + 3’ UTR) https://static-content.springer.com/esm/art%3A10.1038%2Fs41586-025-09272-9/MediaObjects/41586_2025_9272_MOESM10_ESM.xlsx.

## 11 Ethics statement

This study only used data which was previously published and publicly available, as cited in the manuscript. This SwissPedHealth study, under which this work was carried out, was approved based on the advice of the ethics committee of Northwest and Central Switzerland (EKNZ, AO_2022-00018). The study was conducted in accordance with the Declaration of Helsinki.

## 13 Supplemental

### 13.1 Input

**Tables S1** and **S2** demonstrate the minimal input format required by Archipelago, showing the first few lines from validation study 1 (**section 4.1**):

1. The VSAT table (**Table S1**), typical of SKAT, contains one row per tested variant set (e.g. gene or pathway), identified by `set_ID` and associated P value. As VSAT results are aggregated across multiple variants, no genomic position is provided.
2. The GWAS table (**Table S2**), typical of Plink, contains single-variant results, with genomic coordinates (`CHR` and `BP`) and a P value for each SNP. To enable integration, each SNP must also be annotated with a `set_ID`, matching the grouping used in the VSAT layer.

This shared `set_ID` acts as a join key across resolution layers, allowing Archipelago to combine variant associations with aggregated signals from gene, pathway, or other set-based tests. To align SNP-level set-level results, a shared common key might by missing especially from independent studies. In these cases we used R biomaRt to map the consensus feature based on Ensembl GRCh38 genomic coordinates.

**Table S1:**
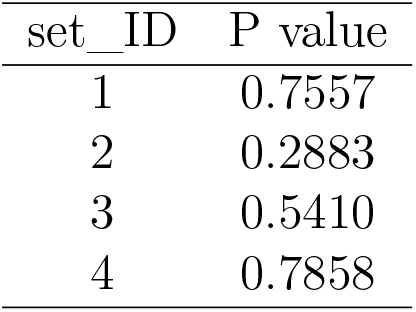
VSAT results from SKAT-O. Each variant set (i.e. protein pathway) requires an index by `set_ID` resulting in a single P value per association test (source: validation study 1KG `df_pathway_sim.csv`).

**Table S2:** GWAS results from Plink. In GWAS, each variant is reported with its chromosome (`CHR`), base-pair position (`BP`), and association P value. To enable integration with the VSAT results in Table S1, each SNP is annotated with its corresponding `set_ID`, derived by mapping to GRCh38 genes grouped into protein pathways. (source: validation study 1KG `df_snp_id.csv`).

| set_ID | CHR | BP (position) | P value |
| --- | --- | --- | --- |
| 1 | 1 | 160289049 | 0.2560 |
| 1 | 1 | 160295829 | 0.5597 |
| 1 | 1 | 160300635 | 0.9210 |

### 13.2 Validation methods

#### 13.2.1 Validation method in 1KG GWAS and simulated gene-level VSAT

We performed a validation test using a popular reference dataset consisting of 504 EAS individuals from 1000 Genomes Project phase 3 version 5 (1KG 3v5), genome build human_g1k_v37.fasta (hg19) (http://ftp.1000genomes.ebi.ac.uk/vol1/ftp/release/20130502/) (31). This dataset contained around 1 million variants from the ancestral backgrounds: CHB: Han Chinese in Beijing, China; JPT: Japanese in Tokyo, Japan; CHS: Southern Han Chinese; CDX: Chinese Dai in Xishuanagbanna, China; and KHV: Kinh in Ho Chi Minh City, Vietna. The result of this study is shown in **Figure 4**.

To streamline reproducibility we processed this dataset using the popular tutorial from the Laboratory of Complex Trait Genomics (Kamatani Lab) in the Deparment of Computational Biology and Medical Sciences at the Univerty of Tokyo (https://github.com/Cloufield/GWASTutorial). The data was processed in preparation for a typical GWAS study: we selected only autosomal variants, split multi-allelic variants, variants were normalized, remove duplicated variants, selected only SNP (ATCG), selected 2% rare SNPs (`plink –mac 2 –max–maf 0.01 –thin 0.02`, selected 15% common SNPs (`plink –maf 0.01 –thin 0.15`), converted to plink bed format and merged to a single file, and randomly added some missing data points. A prepared version of this processed data is also available from the authors repository in zipped Plink format. Since this public data set (1KG 3v5) as no shared disease phenotype, we followed the same resource to simulate the phenotype file. The simulation parameters were: number of simulation replicate(s) = 1 (Default = 1), heritability of liability = 0.8 (Default = 0.1), disease prevalence = 0.5 (Default = 0.1), number of cases = 250, and number of controls = 254. The QC steps included running –r2 with the following filters: `–ld-window: 10, –ld-window-kb: 1000, –ld-window-r2: 0.2`, and `–r2 to plink_results.ld`. We then included `–geno 0.02, –hwe 1e-6, –mind 0.02`. High-LD regions were flagged (`high-ld-hg19.txt`) and pruning was done with `–indep-pairwise 500 50 0.2`. For subject kinship, KING-cutoff 0.0884 was followed by projection (related and unrelated samples).

We performed the single-variant GWAS as a logistic regression with firth correction for a simulated binary trait under the additive mode, with `–maf 0.01`, five PCs, and the simulated phenotypes. Next, we used the default protein pathway gene sets from ProteoMCLustR (https://github.com/DylanLawless/ProteoMCLustR). This consists of a pre-computer set of pathways after cluster the full human genome set of STRINGdb with PPI evidence confidence score 0.7 and contains 941 distinct pathways. We mapped these pathways ID with the variant coordiates (bim file) using the gene coordiates available for this genome build from ensembl biomart `mart_export_GRCh37p13.txt` (https://grch37.ensembl.org/biomart/martview/) for the dataset: human genes (GRCh37.p13), with attributes: Gene stable ID, Gene start (bp), Gene end (bp), Chromosome/scaffold name, Gene name. Our repository also includes `mart_export_GRCh38p14.txt` from http://mart.ensembl.org/biomart/martview/.

We then performed the VSAT with pathway-level variant collapse by running SKAT-O with the `set_ID` assigned to these ProteoMCLustR pathways (e.g. pathway 1,…,n). After QC in GWAS this cohort contained 500 samples and in VSAT we had 945 sets with 1224104 total SNPs. We saved the results from both the Plink single-variant GWAS and the pathway-level SKAT-O. Since this is not a disease cohort and the phenotype is simulated, we automatically increased the association strength of the top VSAT set to provide an enrichment signal. This result is similar to what we have observed in other studies of this size but which do not have public genetic data. Running Archipelago simply requires providing two such inputs (i.e. VSAT results, GWAS results) and any custom theme settings: `archipelago_plot(df_pathway_sim, df_snp_id`).

The enriched variant signals displayed in **Figure 4** were assessed based on their allele frequency. For the GWAS, the allele frequency of enriched variants (n = 67) was: mean = 0.333, median = 0.315, min = 0.133, max = 0.49 which demonstrates that these individual SNP associations were due to highly common variants. For the top VSAT hit (`set_ID` 532), there were 261 variants. The allele frequency of variants was: mean = 0.151, median = 0.121, min = 0.011, max = 0.478. This demonstrates the scenarios where rare variants within a collapsed set can have a different association than the GWAS region, although this is not always true.

**Figure S1:**
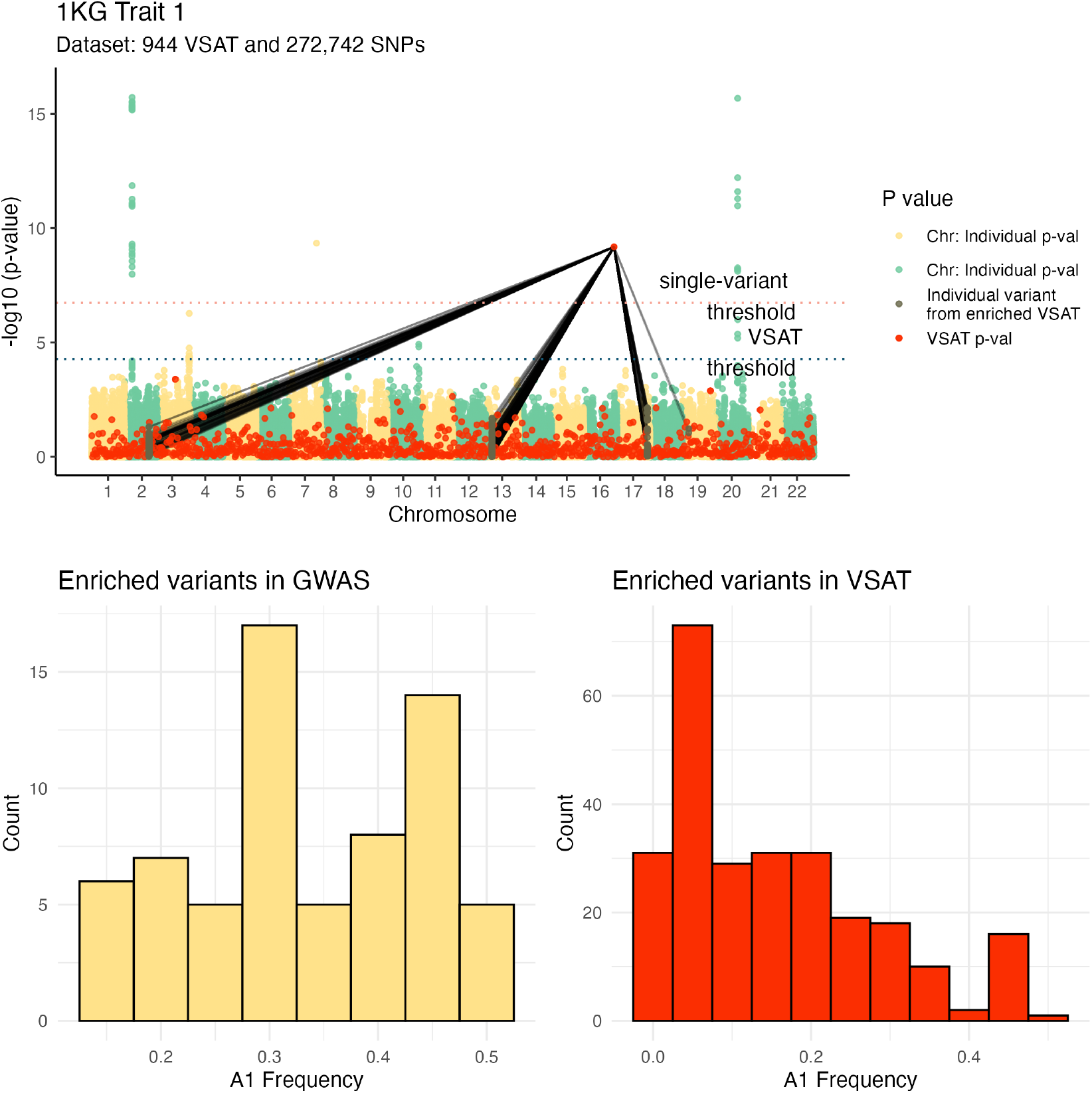
Allele frequency distributions and signal architecture in the 1KG simulation. Top: Archipelago visualisation of SNP-level GWAS and pathway-level VSAT results for the simulated binary trait in the 1000 Genomes East Asian cohort. Bottom: Frequency distributions for enriched variants. GWAS-enriched SNPs had a higher mean allele frequency (mean = 0.33) than the top pathway signal (`set_ID` 532, mean = 0.15), highlighting the complementary resolution of common and rare variant associations.

#### 13.2.2 Validation method in Pan-UK Biobank using GWAS and gene-level DeepRVAT

To evaluate performance on large-scale data, we applied the Archipelago method to the quantitative UK Biobank trait *platelet distribution width* (phenocode 30110), using two layers: the single-variant level GWAS performed by Karczewski et al. (27) and the gene-level RVAT (which is a VSAT focusing on rare variants) performed in DeepRVAT by Clarke et al. (28).

We retrieved GWAS summary statistics from the Pan-UK Biobank resource (27), comprising 29 million SNPs across 472,000 participants. The gene-level VSAT summary statistics were obtained from the DeepRVAT dataset (6), a published resource accompanying the DeepRVAT method (28). DeepRVAT integrates functional annotations via deep set networks to estimate trait-agnostic gene impairment scores. It tested 3.49 million gene-trait pairs across 97 traits using UK Biobank whole-exome sequencing (WES) data.

To align SNP-level GWAS and gene-level RVAT results, we mapped common variants to protein-coding genes using Ensembl GRCh38 genomic coordinates. The harmonised layers used a shared `set_ID`, defined as the gene ID. For genes without overlapping SNPs (*n* = 7), we assigned the first nearest proxy SNP. Most large-scale studies report summary statistics at a single resolution, so comprehensive overlap between independent GWAS and RVAT datasets remains uncommon. In this analysis, prominent common variant GWAS signals were complemented by additional, independent gene-level RVAT associations whose contributing rare variants were not directly represented in the GWAS layer.

For tractability in plotting, we automatically downsampled the 35,968 gene-level RVAT results by using a ramped log-scale weighting based on − log_10_(*p*). This prioritised moderately significant genes while reducing visual clutter from uniformly null signals, yielding a focused subset of 396 genes for plotting. Data were trimmed at P < 1e-75 for visibility due to large outliers. For easier reproducibility, we also included a randomly downsampled GWAS using 1 in 200 of the 29 million SNPs (yielding ~145,000 variants). Had summary statistics been publicly available at the rare variant level we would also expect additional mapping between the two layers.

#### 13.2.3 Validation method in UKBB WGS UTR collapsing PheWAS

We integrated two layers for a binary phenotype: SNP-level GWAS from the PanUKBB resource (27) and gene-level UTR collapsing results from the UKBB WGS PheWAS (8). Here, The UK Biobank Whole-Genome Sequencing Consortium et al. (8) report in Supplementary Table 16 (sheet “Binary”) for the 5’ UTR, 3’ UTR and 5’+3’ UTR models. We provided this dataset as the input for the Archipelago VSAT layer. For GWAS, as previously discussed, we mapped variants to protein-coding genes using Ensembl GRCh38 coordinates. We set the VSAT significance threshold based on the original study’s guidance for PheWAS (*P* ≤ 1 *×* 10^−8^) The UK Biobank Whole-Genome Sequencing Consortium et al. (8) which is equivalent to 5 million tests with Archipelago’s default for Bonferroni correction. Archipelago then visualised the aligned layers and linked set-level UTR associations to the gene-based `set_ID` GWAS variants at the *HBB* locus.

### 13.3 Figure layers

**Figures S2a, S2b**, and **S2c** contain the the same dataset as **Figure 1** however, each version illustrates the decreasing levels of clarification information to demonstrate the layers of annotation. **Figure S2a** shows the original plot without the figure legend. **Figure S2b** drops the edge highlights for the significantly enriched VSAT to reveal all connections, which can be difficult to read in high density plots. **Figure S2c** next drops the individual variant P value highlighting.

**Figure S2:**
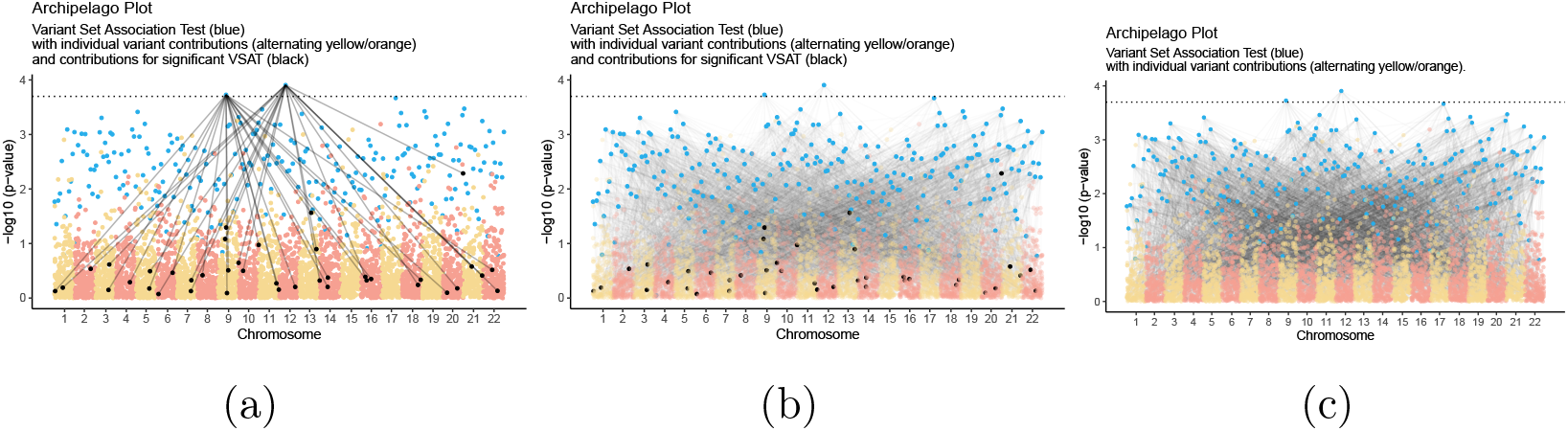
Illustration of information layers used to reduce complexity.

### 13.4 Variant set edge colour

**Figure S3** adds an additional layer of information to that seen in **Figure S2a** by adding two colours for the significantly enriched variant sets which is useful for when there are multiple enriched variant sets but otherwise may be distracting.

**Figure S3:**
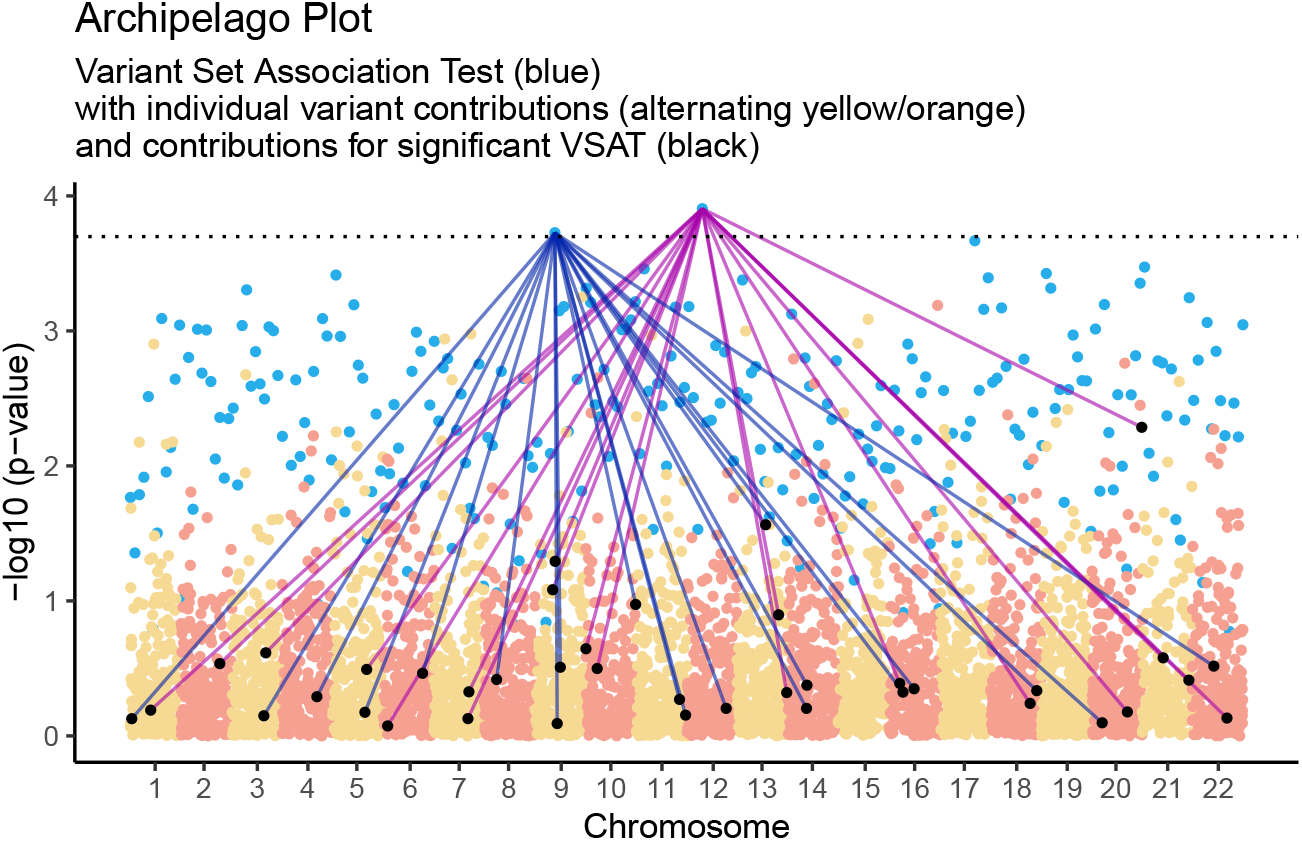
Colored VSAT edges to indicate separate variant sets from two significantly enriched VSAT P values.

### 13.5 Sparse plots

We demonstrate a set of sparse plots in **Figures S4a, S4b**, and **S4c**. These smaller dataset examples use synthetic data to represent 500 qualifying variants, and VSAT of 20 genes/variants per variant set (25 VSAT P values). **Figure S4a** illustrates a small variant set of 500 individual variants in 25 variant sets. **Figure S4b** and **Figure S4c** continue by dropping information layers sequentially as previously described in the dense plot examples.

**Figure S4:**
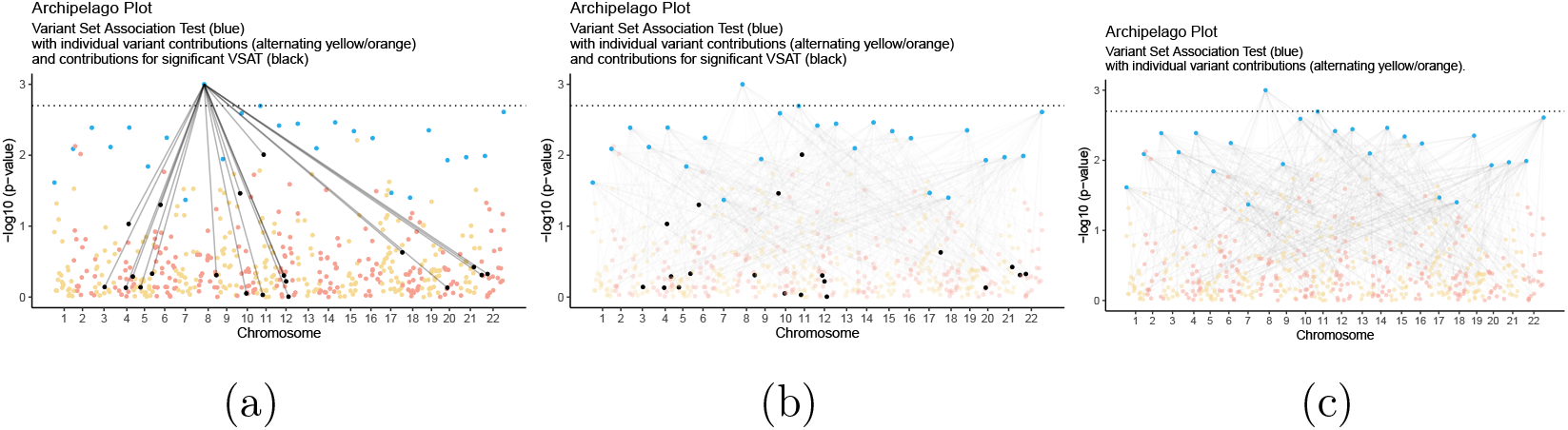
Illustration of information layers used to reduce complexity in a sparse dataset.

### 13.6 R package user settings

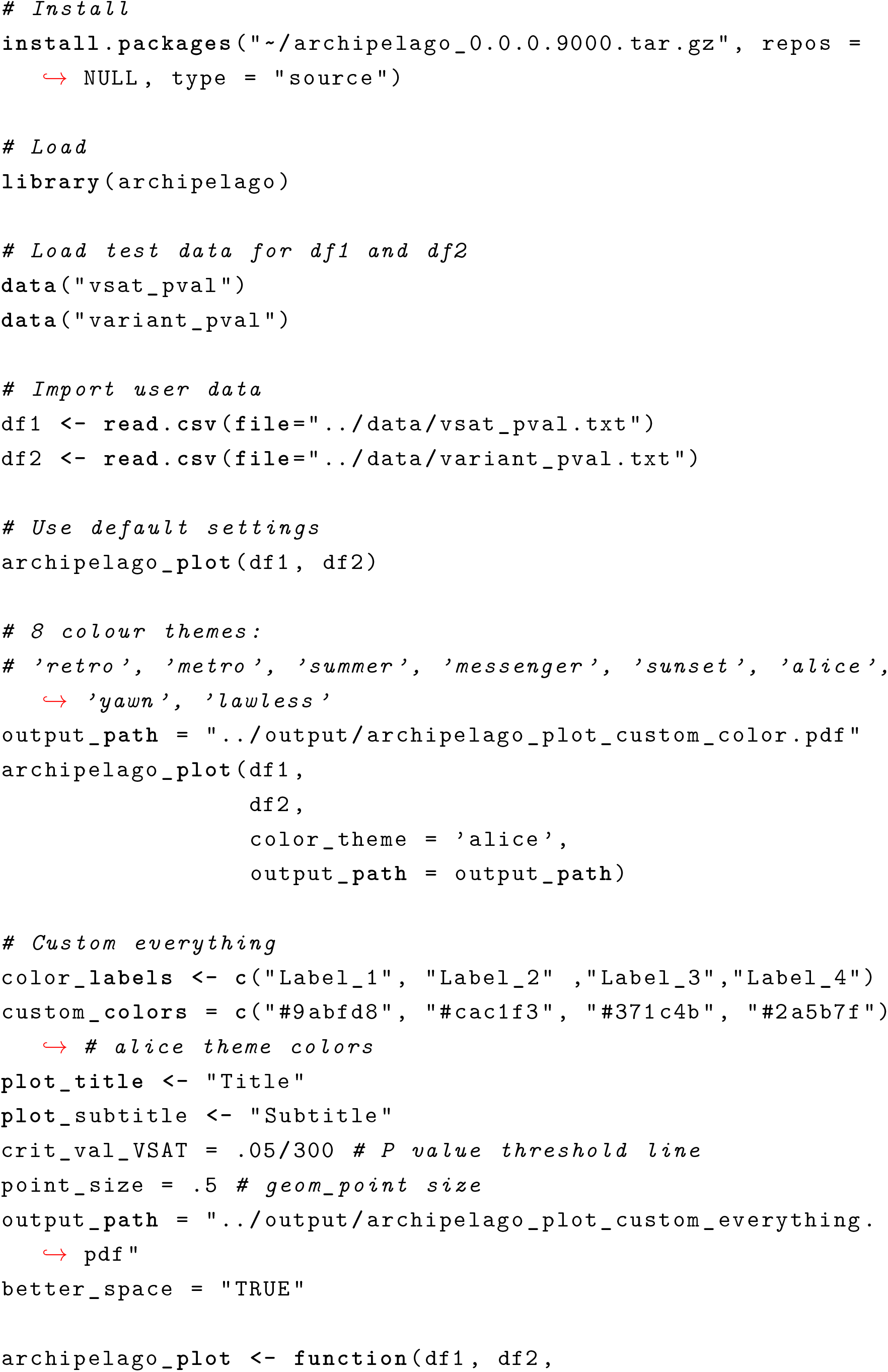

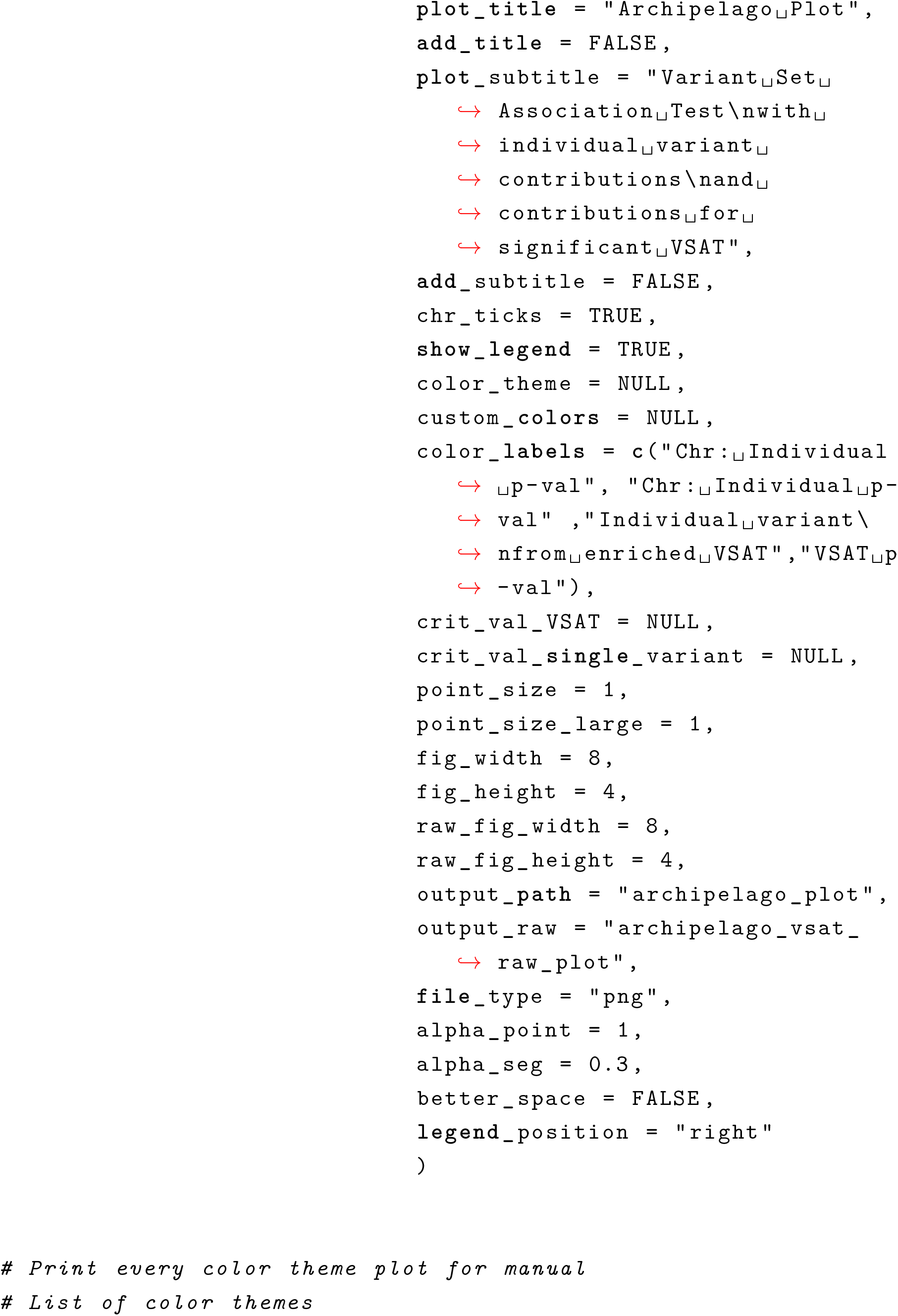

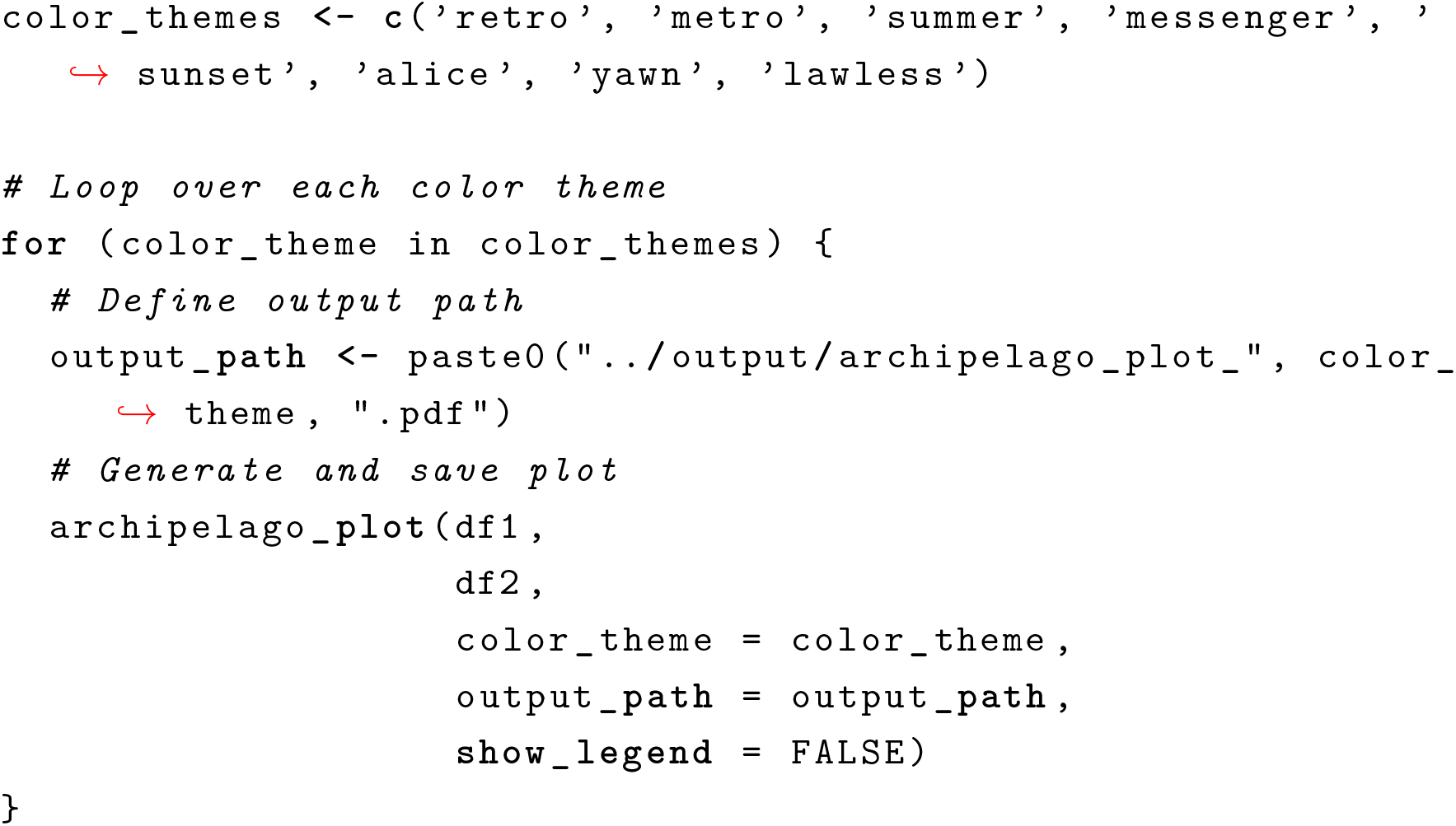

### 13.7 Protocol Summary

The Archipelago Plot protocol takes as input two datasets containing variant-set association testing (VSAT) P values and single variant P values. It first orders the variant P values by chromosome and genomic coordinate, then averages the coordinates within each variant set. The protocol subsequently normalizes the VSAT x-axis distribution (optional) to avoid center clustering in dense datasets, prioritizing VSAT P values in case of overlaps. It can optionally map the VSAT position to each variant from the set. The protocol then creates a ggplot visualisation, which includes an option to highlight individual variant contributions from significant VSATs.

### 13.8 Protocol Formal Definition

The formal definition of the Archipelago Plot Protocol comprises the following steps: **Data Preparation**: The data, including variant-set association testing (VSAT) P values and single variant P values, is sorted and averaged within each variant set. **Normalization**: Optionally, the VSAT x-axis distribution is normalized to avoid center clustering in dense datasets. Ranking is done to prioritize VSAT P values in the event of overlaps. **Mapping**: Optionally, the position of each VSAT is mapped to each variant from the set. **Visualisation**: A plot is created using ggplot2, which includes options to add a title, subtitle, chromosome ticks, a legend, and custom colours. **Highlighting**: The plot can be made to highlight individual variant contributions from significant VSATs. **Output**: The plot is saved to a specified path and returned by the function.

### 13.9 Protocol Algorithm

The core of the algorithm can be broken down into the following steps: **Pre-processing**: Load required libraries (ggplot2 and dplyr). Determine the critical value if it’s not provided. Data ordering and coordinate calculation: Sort the single variant P values by chromosome and genomic coordinate, and create an index to order the chromosomes. Calculate the position for each data point, essentially ordering the chromosomes end to end, and create mid-points for each chromosome for labeling. **Merging and preparation**: Merge the VSAT P values with the single variant P values, then calculate the sum and average position of each variant set group. Replace missing positions with the average position of their variant set. **Normalization**: If chosen, normalize the VSAT x-axis distribution. This is done by sorting the variant set P values by their position, then assigning evenly spaced numbers across the range of all position values. **Data colouring and splitting**: Assign colours to the different groups in the data and split the data into variant sets and individual variants. **Highlighting**: Define a condition to highlight individual variant contributions from significant VSATs and apply it to the data. **Plotting**: Create the plot with ggplot2, add colour scaling, labels, theme, and optionally title, subtitle, and legend. **Finalization**: Save the plot to a specified output path and return the plot from the function.

The R code is relatively easy to modify or rewrite. Alternative versions could be written based on the following algorithm:

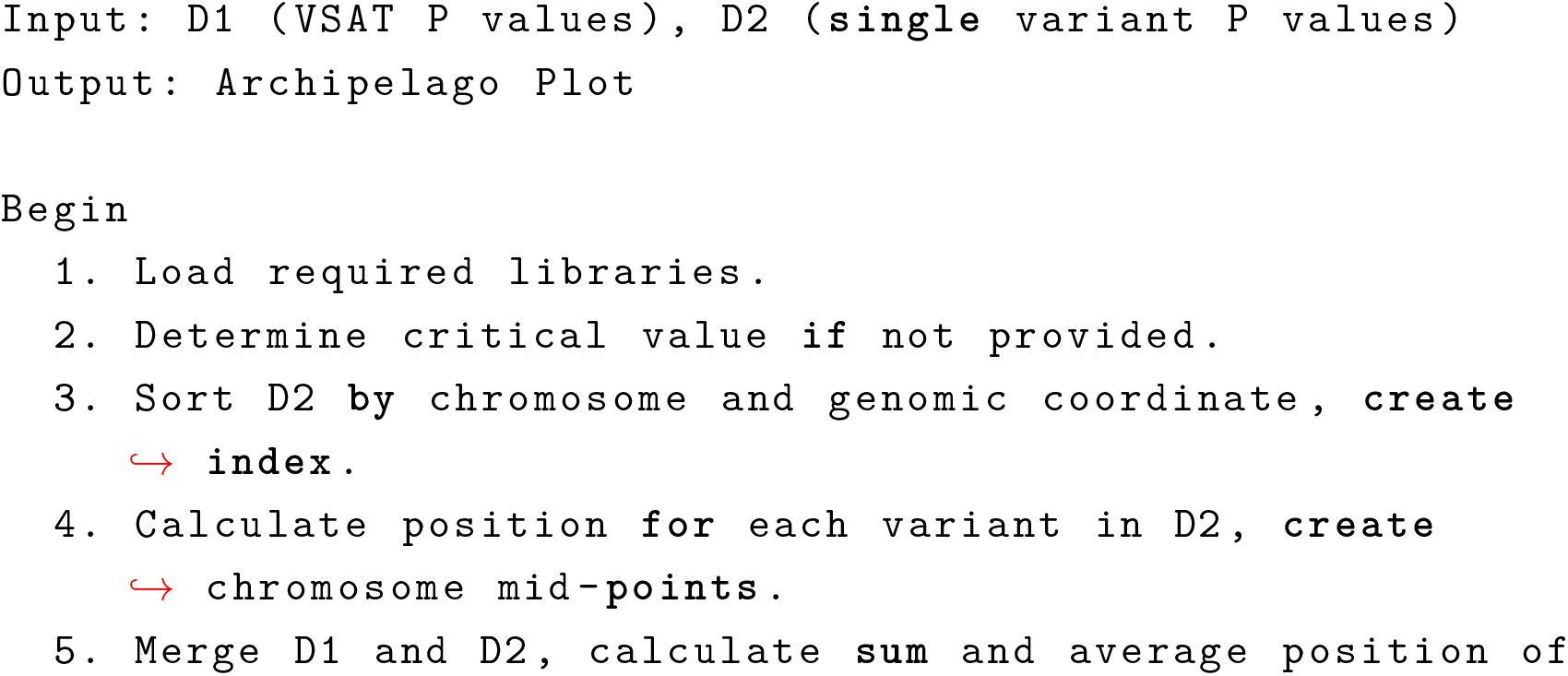

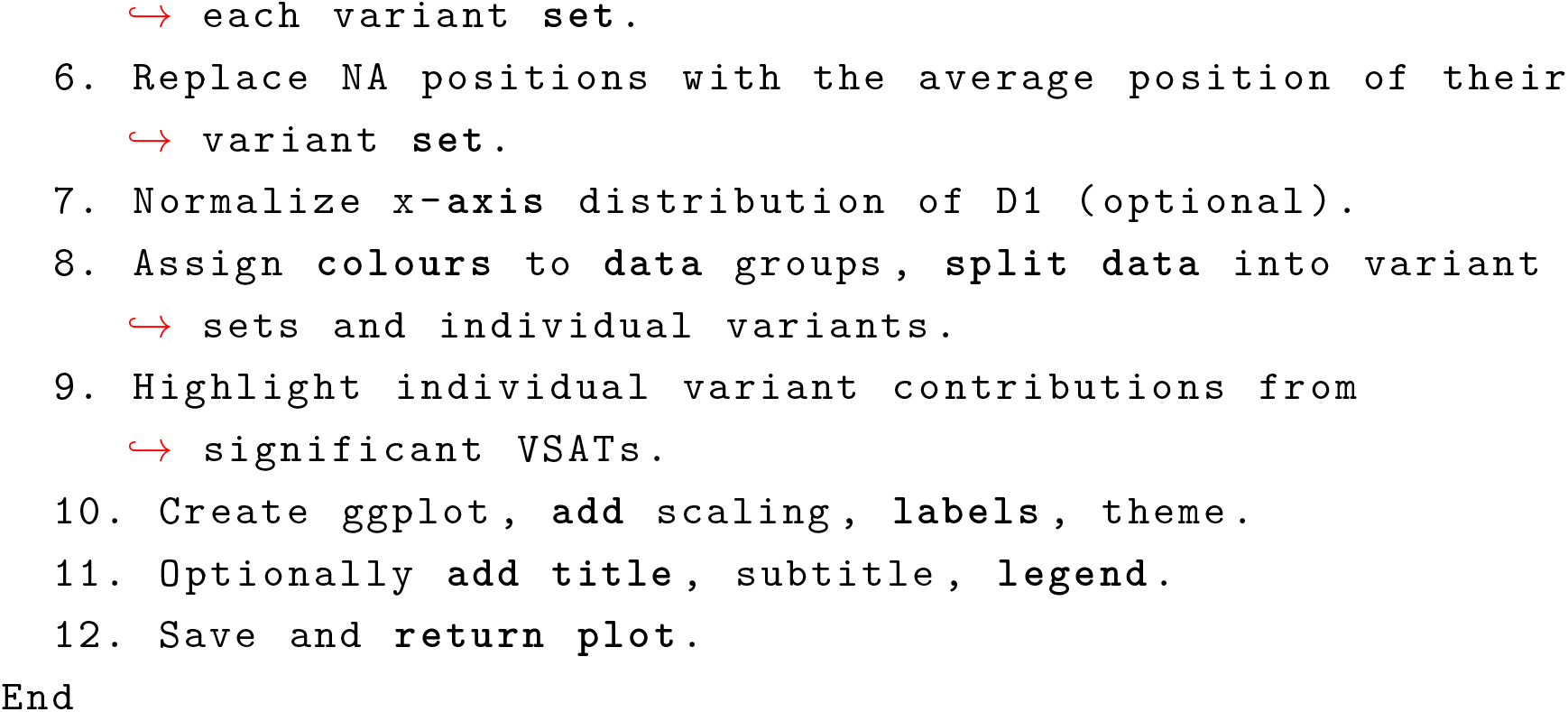

### 13.10 Customisation

Figure S5 shows an example of customised colours. Figure S6 shows all customisable elements. Figure S7 - S8 shows all 16 colour themes.

**Figure S5:**
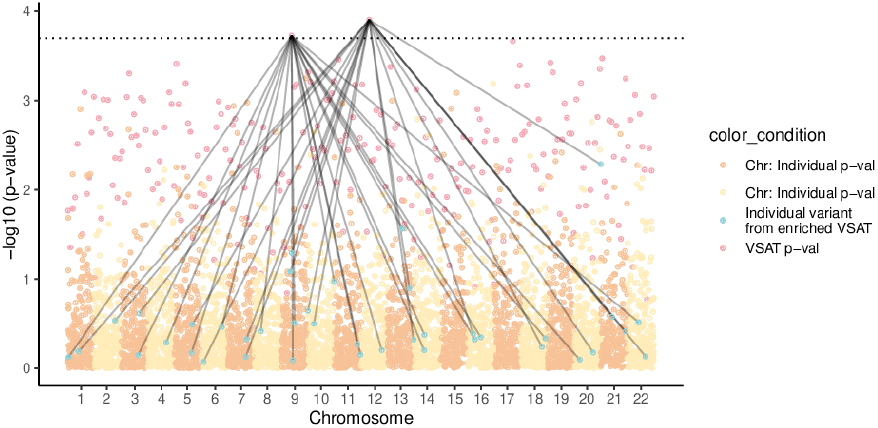
Custom colours specified by hex values, colour names, etc.

**Figure S6:**
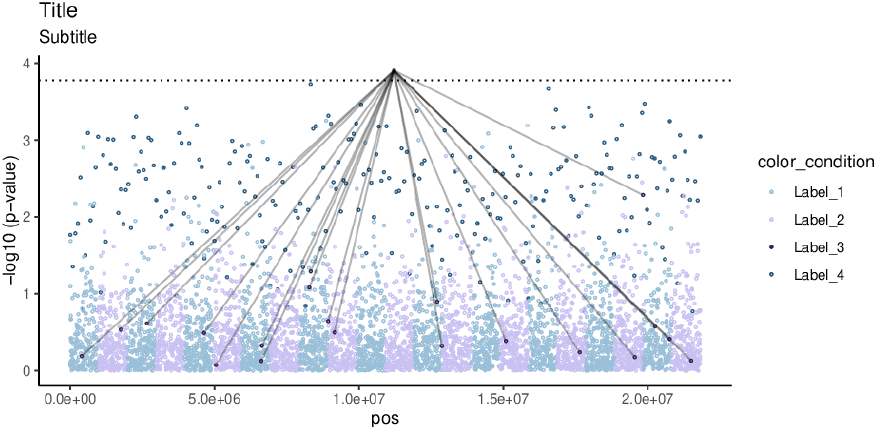
Custom title, subtitle, colours, colour labels, critical threshold line, genomic coordinate, show title and subtitle, and show legend.

**Figure S7:**
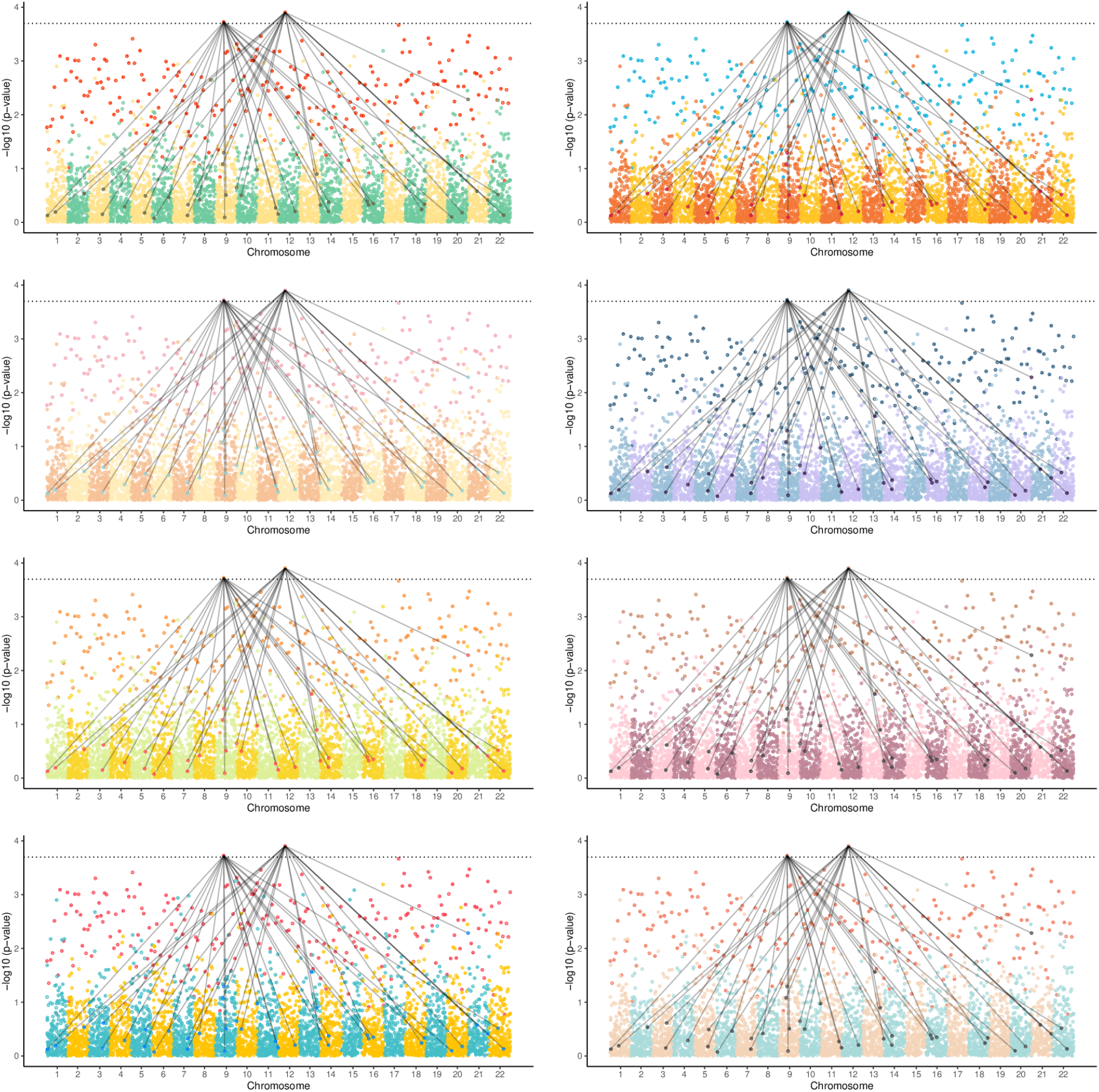
Color themes left to right: ‘retro’, ‘metro’, ‘alice’, ‘buckley’, ‘summer’, ‘romance’, ‘messenger’, ‘meme’.

**Figure S8:**
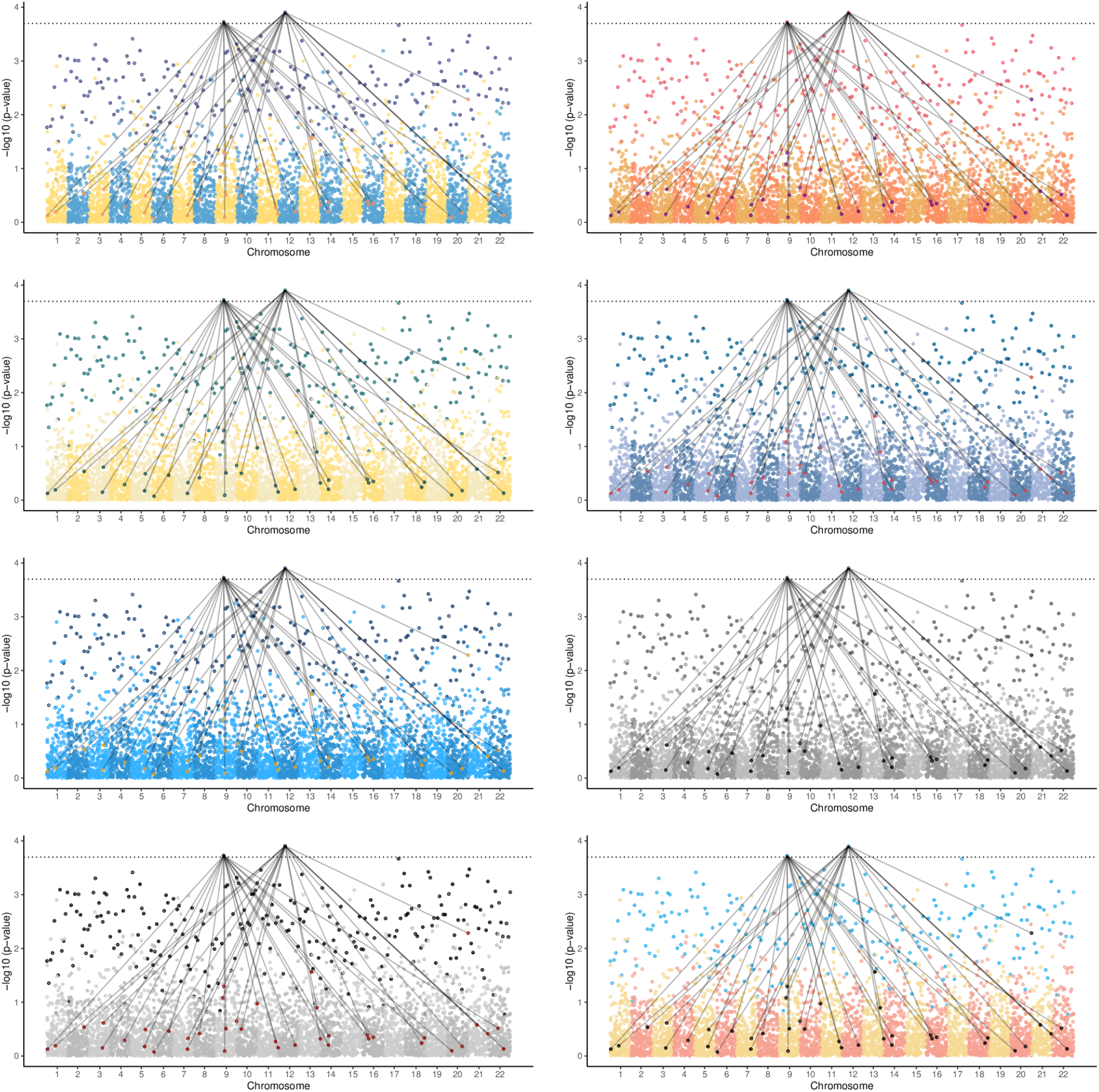
Color themes left to right: ‘pagliacci’, ‘sunset’, ‘ambush’, ‘saiko’, ‘sunra’, ‘yawn’, ‘caliber’, ‘lawless’.

**Figure S9:**
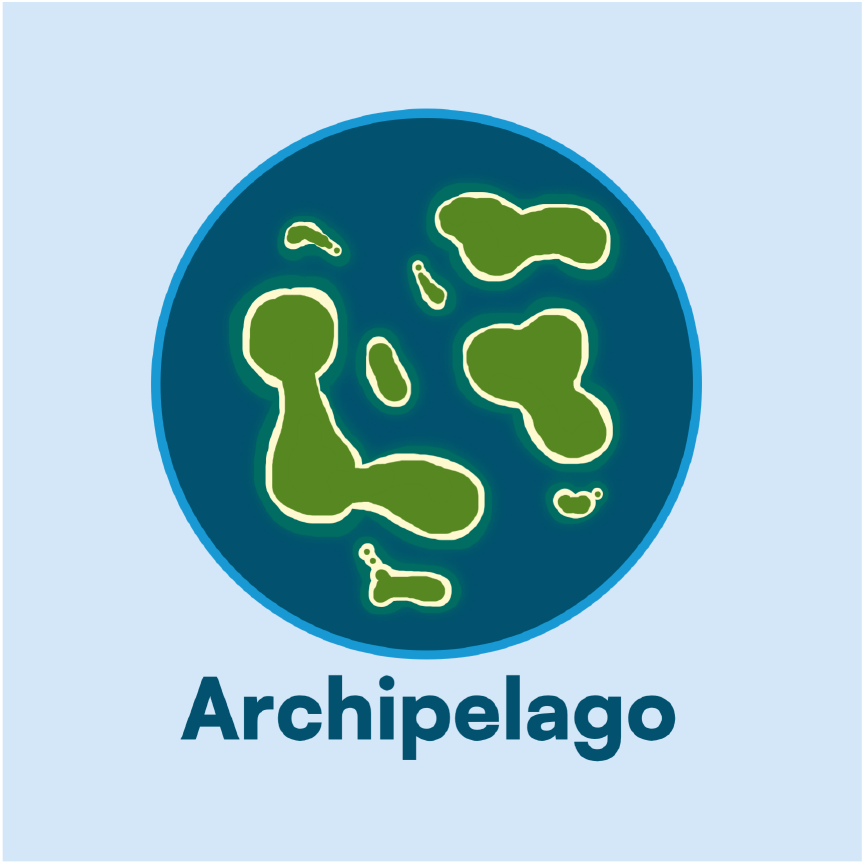
Archipelago plot logo.

